# Gene prioritization in GWAS loci using multimodal evidence

**DOI:** 10.1101/2023.12.23.23300360

**Authors:** Marijn Schipper, Christiaan A. de Leeuw, Bernardo A.P.C. Maciel, Douglas P. Wightman, Nikki Hubers, Dorret I. Boomsma, Michael C. O’Donovan, Danielle Posthuma

## Abstract

Genome-wide association studies (GWAS) yield large numbers of genetic loci associated with traits and diseases. Predicting the effector genes that mediate these locus associations remains challenging. Here we present the FLAMES framework, which predicts the most likely effector gene in a locus. FLAMES integrates machine learning predictions from biological data linking single nucleotide polymorphisms (SNPs) to genes with GWAS-wide convergence of gene interactions. We benchmark FLAMES on gene-locus pairs derived by expert curation, rare variant implication, and domain knowledge of molecular traits. We demonstrate that combining SNP-based and convergence-based modalities outperforms prioritization strategies using a single line of evidence. Applying FLAMES, we resolve the *FSHB* locus in the GWAS for dizygotic twinning and further leverage this framework to find novel schizophrenia risk genes that converge with rare coding evidence and are relevant in different stages of life.

## INTRODUCTION

Genome-wide association studies (GWAS) have revolutionized modern genetics by providing a data-driven approach to discovering genetic factors involved in complex traits. Large sample sizes in GWAS have resulted in thousands of trait-associated single nucleotide polymorphisms (SNPs). These associations between SNPs and traits can provide valuable insight into the underlying biological mechanisms. However, in the case of polygenic phenotypes with which hundreds of SNPs with small effects are associated, the phenotypic impact of a single SNP may be difficult to interpret. As genes are the universal effectors of biological processes, it is assumed that the effects of most trait-associated SNPs arise from SNPs impacting gene regulation or gene products. Translating SNP-trait associations to gene-trait associations allows us to place GWAS findings in a broader biological context.

Recently, integrative methods have been developed that aim to combine many different levels of functional data to predict the effector gene in a GWAS locus^1–4^. There are two main strategies to do so. The first strategy prioritizes genes using locus-based SNP-to-gene data. Examples of this are chromatin interaction mapping, QTL mapping, or selecting the closest gene to the lead variant. These annotations can then be combined by linear regression or machine learning to merge multiple types of SNP-to-gene data into a single prediction^1,3^. The second strategy assumes that all GWAS signal converges into biological pathways and networks. These methods prioritize genes in a locus based on gene features enriched across the entire GWAS^4^. However, no current method leverages these two strategies together to make well-calibrated predictions of the effector genes in a locus^5^.

We designed a new framework, called Fine-mapped Locus Assessment Model of Effector geneS (FLAMES). This framework integrates SNP-to-gene evidence and convergence-based evidence into a single prediction for each fine-mapped GWAS signal. We benchmark the performance of our method against current state-of-the-art methods for three well-understood molecular traits^6^, an expert-curated gold standard set^3^, and ExWAS results of nine traits. We show that FLAMES outperforms approaches that leverage only a single strategy, and that our integrative method accurately predicts effector genes from GWAS data. We leverage this new method to establish a suspected link between *FSHB* and giving birth to dizygotic twins and find novel temporally relevant gene sets related to schizophrenia.

## RESULTS

### A framework for combining evidence linking SNPs to genes and biological convergence from fine-mapped GWAS loci

We created an integrative framework called Fine-mapped Locus Assessment Model of Effector geneS (FLAMES), which combines SNP-to-gene evidence and convergence-based evidence, outputting a single score per gene (Fig. 1). In summary, this framework annotates fine-mapped credible sets and uses a machine learning classifier to score each gene, where this score denotes the level of biological evidence for that gene being regulated by a set of credible causal SNPs in the locus. The exact annotations used to generate the biological evidence can be found in table 1. The XGBoost^7^ classifier used to create the SNP-to-gene scores is trained on a set of GWAS loci that contain a gene implicated by predicted loss of function (pLoF) variants or missense variants associated with the corresponding trait in an exome-wide association study (ExWAS). The scores are subsequently combined with convergence-based PoPS scores^4^. This results in a single FLAMES score per gene in the region for each fine-mapped GWAS signal. The assumption underlying the FLAMES framework is that for each true GWAS association between SNP and phenotype that is not due to population stratification or other form of confounding, there is a gene that most strongly mediates the effect of the SNP on the phenotype. We call the gene that mediates the SNP’s effect on the phenotype the effector gene of said SNP. Therefore, each credible set in a locus should have an effector gene. FLAMES aims to predict a single most likely effector gene for each credible set, and will output a gene scores for each separate credible set. The FLAMES score of a gene denotes the combined evidence of functional convergence with other GWAS implicated genes and the evidence from biological studies based on the GWAS SNPs in the locus of interest.

**Figure 1.**
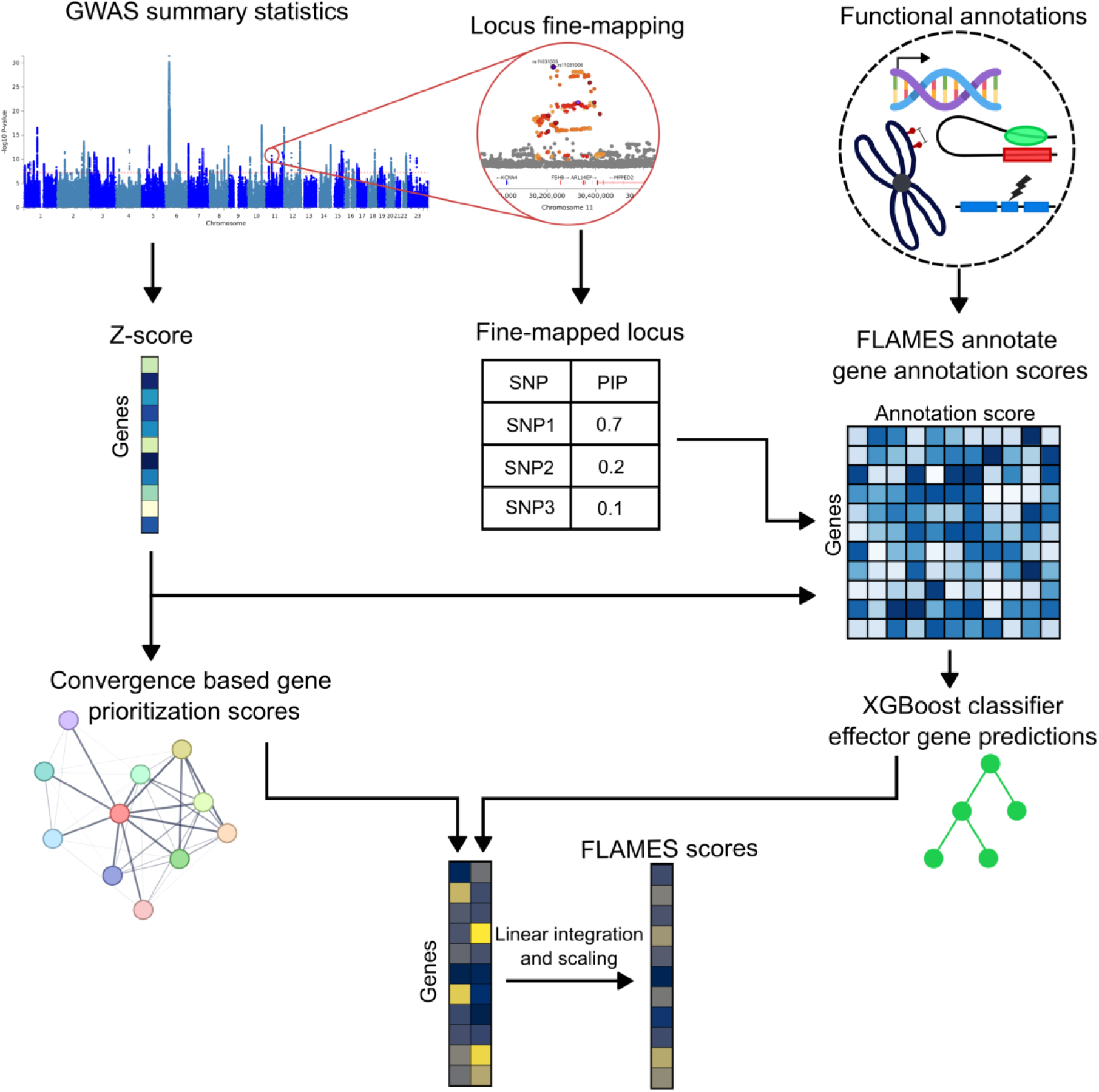
FLAMES framework. Overview of the FLAMES framework. Functional annotations scores are calculated by integrating fine-mapped credible sets with SNP-to-gene evidence. Biological data used to create annotation scores can be found in Table 1. Gene Z-scores are derived using MAGMA^8^. Convergence based gene prioritization scores are calculated with PoPS^4^. The XGBoost classifier used to create gene scores is described in the methods.

**Table 1.**
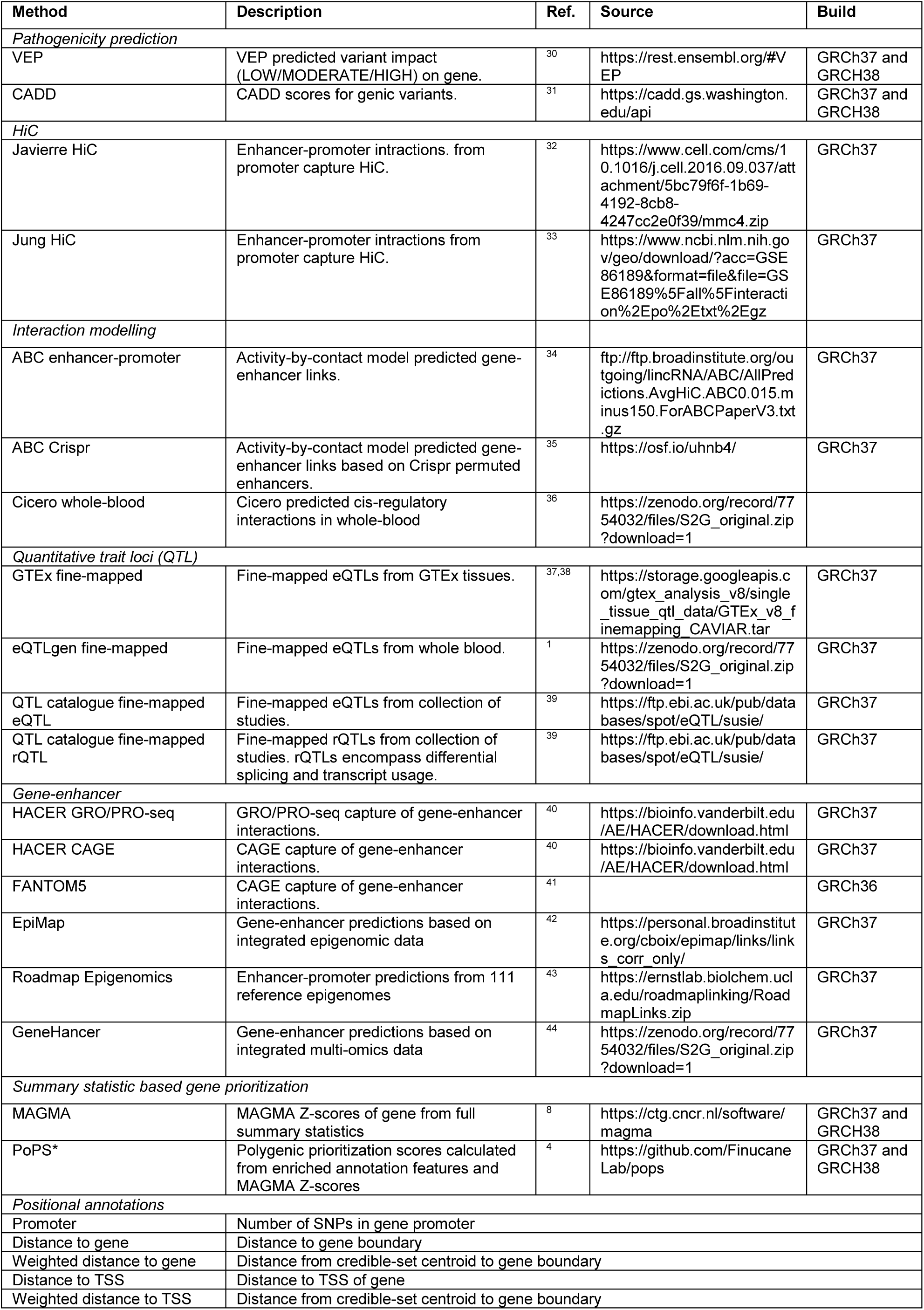
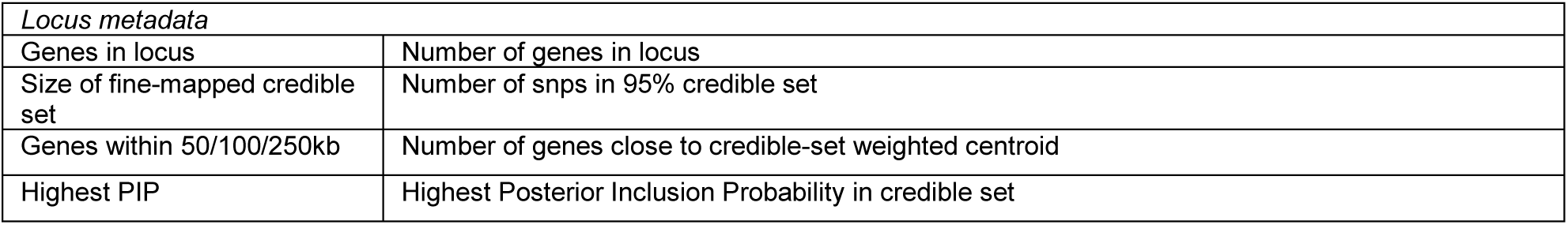
Functional annotations integrated in FLAMES. Download file of integrated raw data provided in the source column. PoPS is not used in the machine learning classifier.

### Predicting effector genes using annotations linking SNPs to genes via a data-driven approach

When creating a model that prioritizes genes based on SNP-to-gene annotations, it is crucial that the ground truth on which the model is trained represents the general biology underlying GWAS variants as closely as possible in order to achieve optimal generalizability. We therefore hypothesized that a data-driven approach to train a model would result in the highest generalizability. To test this hypothesis, we created a data-driven training set on which we trained FLAMES, and benchmarked FLAMES on multiple distinct datasets against other gene prioritization methods. An overview of the design of this study is given in figure 2. This data-driven training set was created by analyzing 376 heritable traits form the UK biobank (UKBB)^9,10^, and extracting each locus for which a single gene was also significantly associated with the same trait by rare coding missense or predicted loss of function (pLoF) variants in an exome wide association study (ExWAS)^11^. If such an overlap between GWAS common variants and ExWAS rare deleterious variants associated with the same phenotype is found, then these two signals are very likely to be linked to the same gene, and this overlap does not hinge on prior knowledge that might introduce strong biases towards certain annotations. These loci with a rare coding ExWAS association were fine-mapped with FINEMAP^12^ and an in-sample LD reference panel and max one credible set per locus. To annotate these loci we developed FLAMES annotate (methods), a method for annotating credible sets with locus metadata and 22 different SNP-to-gene linking methods (Table 1). Each annotation is given a score that represents the combined probability of a SNP in a credible set being causal and the strength of the evidence linking that SNP to a gene. Since multiple SNPs in the credible set can map to a gene, a mean and maximum score are created for each annotation. A locus scaled annotation is created by linearly scaling the scores so that the highest annotation score equals one and the lowest is equals zero. We subsequently retained loci with just a single ExWAS implicated gene in the locus that is within 750kb of the fine-mapped SNP with the highest posterior inclusion probability (PIP). This resulted in a data-driven set of 1181 fine-mapped loci with an ExWAS implicated effector gene and extensive SNP-to-gene annotations for SNPs in the credible set.

**Figure 2.**
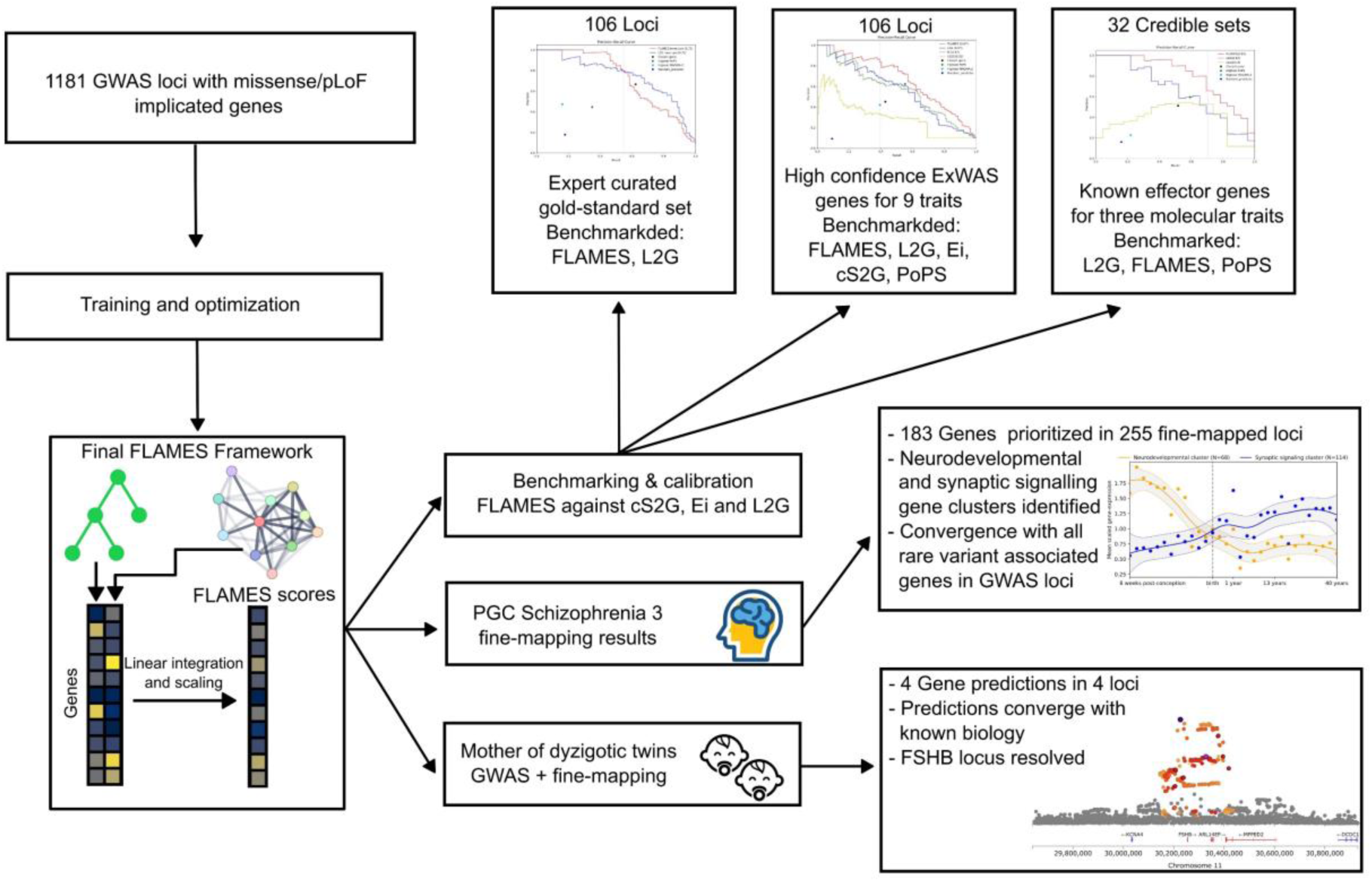
Study design. Overview of study design. 1181 GWAS loci are used to train and optimize a prediction framework which aims to predict the causal gene at a GWAS locus. Benchmarking and calibration are performed on three datasets which the model has not seen before. The model was then applied to a GWAS of being the mother of dizygotic twins and a GWAS of schizophrenia.

To assess whether our data-driven approach captures the genes relevant to our GWAS signals we investigated enrichments of the annotations included in FLAMES annotate. For each annotation we calculated the mean ratio of the ExWAS implicated genes vs the non-ExWAS-implicated genes in the loci, as well as the odds ratio for an ExWAS implicated gene having the highest annotation score in the locus. All annotations are significantly enriched in ExWAS-implicated genes, with the exception for the mean annotation scores of FANTHOM5 enhancer-promoter correlations (Fig 3.a,b, Supplementary Table 2-6). Given the observed positive odds ratios for all annotations, we trained an XGBoost gradient boosting classifier which aims to predict the ExWAS-implicated effector gene for each locus-gene pair from the annotations created with FLAMES annotate on 1181 loci-gene pairs. We assess the importance of the SNP-to-gene annotations using Shapley additive explanations (SHAP) values (Fig. 3.c)^13^. We observe that the overall magnitude of SHAP values resembles the magnitude of odds ratios of ExWAS vs non-ExWAS implicated genes. Generally, high annotation scores are positive predictors across the entire model, which is as expected. Locus meta-data annotations seem to positively impact predictions when the score is low (e.g. few genes in locus, or a small credible set). Overall, the model appears to have integrated the multiple weak predictors into a single model. Given that we observe minimal instances of unexpected negative prediction associated with high annotation scores it is likely that the model has not overfitted to the training data. We subsequently optimized prediction by the integration of PoPS scores using a 50-fold cross-validation approach (see methods). This resulted in a final optimized FLAMES framework where the locus-scaled XGBoost prediction scores and locus-scaled PoPS scores are linearly combined, where XGBoost scores contribute 0.725% and PoPS scores contribute 0.275% to the final FLAMES score per gene. Linear integration of the convergence and SNP-to-gene based methods was chosen to maintain interpretability of the results of the combined methods. To summarize, we created a prediction model that combines SNP-to-gene annotations and convergence-based evidence in order to predict the effector gene of a fine-mapped GWAS locus.

**Figure 3.**
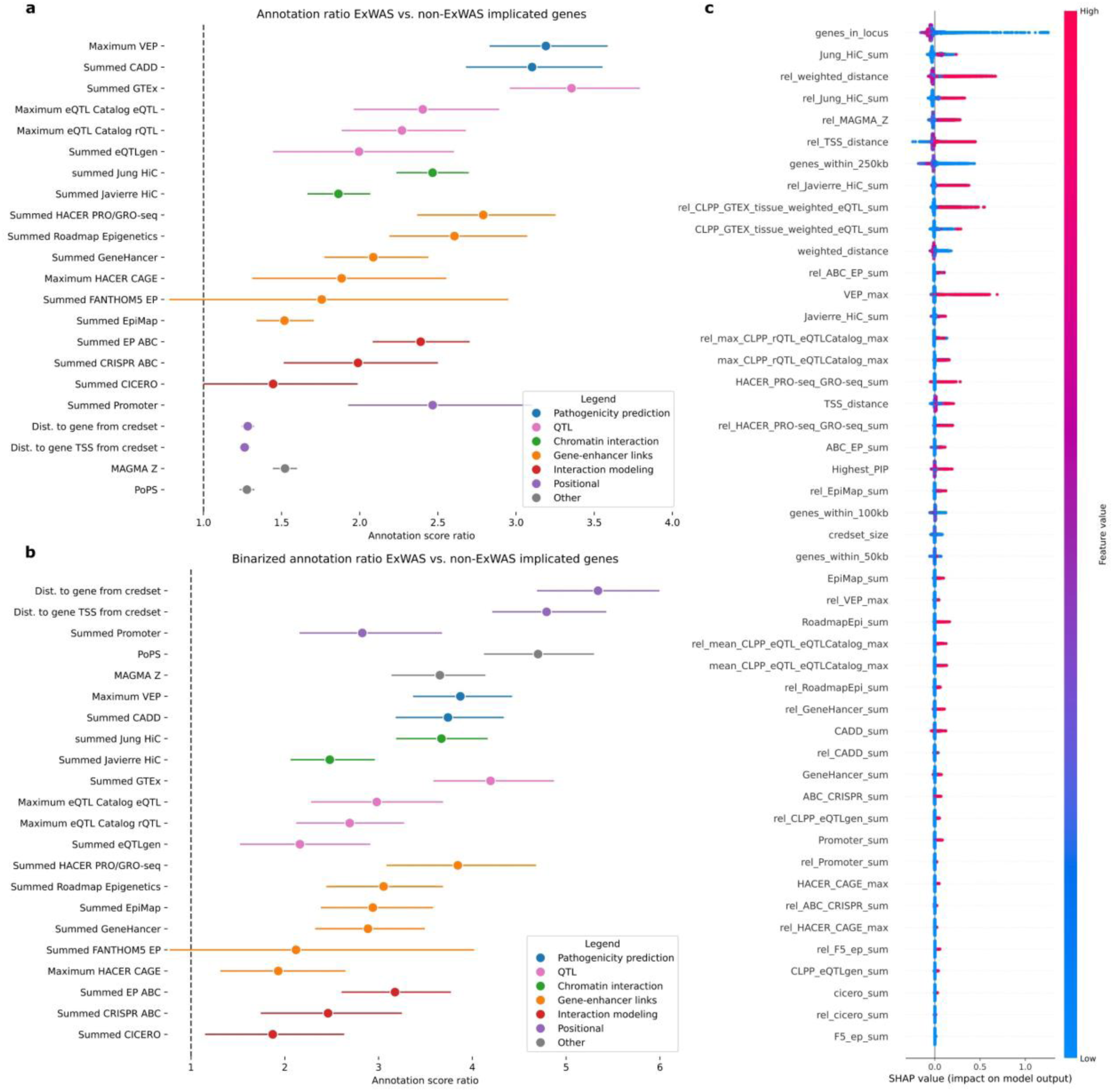
Predictive value of SNP-to-gene annotations. Predictive power of SNP-to-gene annotations assessed in a data-driven ground truth set. **a**, Ratios of average annotation score per annotation in ExWAS implicated gene in GWAS locus vs. the rest of the genes in the locus. Confidence intervals calculated by 1000 times bootstrapping. **b**, Odds ratio of highest annotation score in the GWAS locus belonging to the ExWAS implicated gene in the locus. Confidence intervals calculated by 1000 times bootstrapping **c**, Feature impact on effector gene prediction in XGBoost model. Each feature created by FLAMES is more thoroughly described in Supplementary Table 1.

We repeated the analysis of the predictive value of annotations with an expert curated locus-gene dataset^3^ and compared this to our data-driven locus-gene dataset. We included only loci for which the full GWAS summary statistics were available. We observe that the mean ratios and odds ratios show a significantly stronger enrichment in the expert curated set (Supplementary Figs 1-2). This could be caused by a bias in the selection of locus-gene pairs, purity of fine-mapped credible sets or in the ratio of true to false positives in the locus-gene pairs. Notably, we observe that the odds ratio of PoPS predictions ranks significantly lower for the prediction in the expert curated set compared to the data-driven set. Given that the PoPS predictions are independent of the SNP-to-gene evidence, this implies that there are strong biases in the expert-curated datasets towards certain SNP-to-gene annotations. This is likely due to a selection bias in the curation of these gene-locus-pairs. Overall, the enrichments in annotations implicate that our data-driven approach does indeed capture GWAS relevant locus-gene pairs but avoids biases that can be introduced through expert curation.

### Benchmark of FLAMES

We previously established that our data-driven approach may reduce biases, but could also lead to an increase of mislabeled effector genes in the training dataset. We also hypothesized that our model should generalize well to the datasets outside of the training data if we indeed eliminated selection biases. In order to verify the generalizability of FLAMES we tested our method on 3 datasets that do not overlap with the training data. We benchmarked the performance of FLAMES and other state-of-the-art prediction methods an expert-curated gold standard set, a set of stringently filtered EXWAS predicted genes for nine different traits, and on fine-mapped loci from three interpretable biological traits; serum levels of IGF1, testosterone and urate^6^. Given that Ei^2^, FLAMES and L2G^3^ have different ways of annotating genes to loci, we removed all genes that weren’t annotated by all three methods. cS2G scores were included if they mapped to any gene within the included list. To assess performance we calculated area under the Precision-Recall curve (AUPRC) scores for each method, given that this is the most informative metric in unbalanced dataset. Recall definitions are the same for all benchmarks except for the benchmark on three molecular traits (see methods and Supplementary Note). In this benchmark FLAMES outperforms all other methods, with the exception of L2G on the expert curated (Fig. 4a-c). Notably, L2G precision is likely overestimated as it is predicting on its own training data. To find a recommended threshold for high confidence FLAMES predictions, we combined the results from the dataset of nine traits with ExWAS implicated genes and the dataset of three molecular trait. We then stepwise increased the FLAMES threshold from 0 by 0.05 until we achieved a precision of at least 75%. This resulted in a recommended threshold of keeping FLAMES scores of at least 0.05, the performance at this threshold is visualized in Fig. 4d,e. When using FLAMES at the recommended threshold we see that it outperforms methods that only use SNP-to-gene based evidence or convergence based evidence.

**Figure 4.**
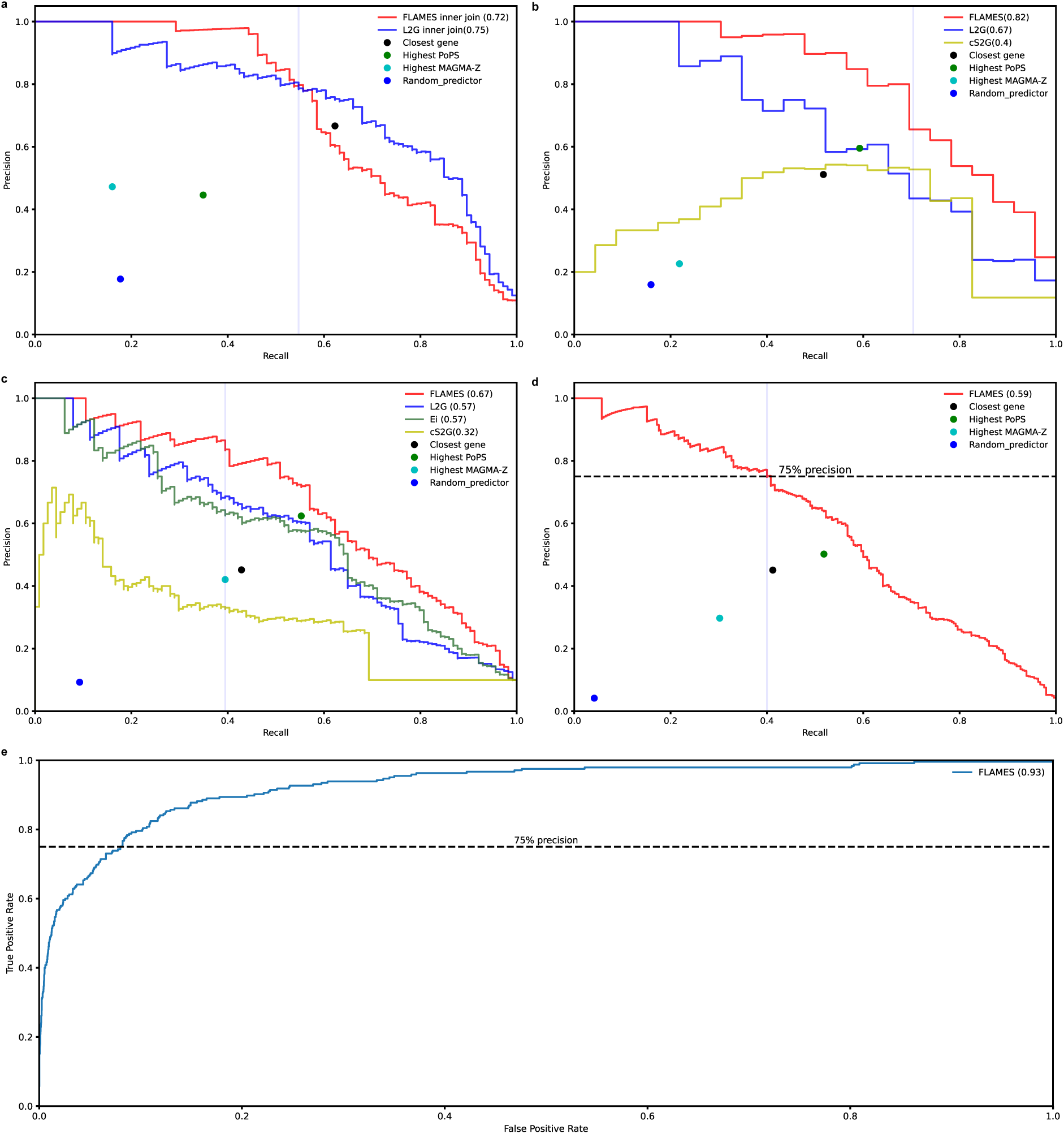
Benchmarking of FLAMES versus state-of-the-art tools in different datasets. Precision-recall curves of different benchmarking sets. Definition of recall may change dependent on the benchmarking set (supplementary notes). Closest gene is reported as the precision and recall of taking the closest gene to the fine-mapped credible sets. In case of ties, all genes are prioritized. Highest PoPS is calculated as the precision and recall of taking the FLAMES annotated gene with the highest PoPS score in the locus. FLAMES threshold of 0.05 is depicted by the blue vertical line. **a**, Precision-recall curve for the performance of FLAMES and L2G on fine-mapped L2G training loci. **b,** Benchmarking of interpretable loci for GWAS of urate, IGF-1 levels and testosterone levels in blood^6^. **c,** Benchmarking of cS2G, Ei, FLAMES and L2G for nine traits^2^. **d,** Calibration of FLAMES threshold in 12 traits combined datases (plot b and c data). The 75% precision threshold corresponds with a FLAMES threshold of 0.05. **e**, Receiver operator curve of FLAMES predictions on plotted in 12 traits combined datasets (plot b and c data)

### Loci for dizygotic twinning GWAS elucidated by FLAMES

We applied FLAMES to a recently completed GWAS of being the mother of DZ twins (MoDZT)^14^. We used FUMA to define loci, which resulted in five distinct loci. We fine-mapped the loci using FINEMAP and a reference panel made up of 100,000 unrelated UKBB individuals, restricting to 1 causal SNP per locus in order to reduce false positives by the out-of-sample reference panel. Given that the original study^15^ reports a merged version of two of our FUMA defined loci we excluded one locus from analysis for consistency with the original GWAS. We retained the locus with the credible set that has the highest Bayes factor, which is the locus that harbors the most likely actual causal variant of the combined locus. In the four different loci, FLAMES prioritizes *GNRH1*, *FSHB*, *SMAD3* and *ZFPM1* for the loci respectively (Supplementary Table 7a,b). *GNRH1* is predicted to be the effector gene of its locus, even though there is a large amount of SNP-to-gene data pointing to *DOCK5* as the causal gene in the locus. *GNRH1* is also the most likely causal gene given its roles in the regulation of follicle-stimulating hormone, luteinizing hormone, and fertility ^16^. The *SMAD3* gene has previously been implicated in MoDZT, although evidence in cattle suggests that *SMAD6* might be the responsible gene^17^. FLAMES suggest both strong functional evidence and convergence-based evidence for *SMAD3*. For SMAD6 there is very little convergence-based evidence for *SMAD6*, and the distance between the fine-mapped SNPs and *SMAD6* is too large for it to be annotated with any SNP-to-gene data. The prediction of *FSHB* in locus 2 is in line with the expected underlying biology of the trait, but is remarkable given the amount of SNP-to-gene evidence linking the fine-mapped SNPs to *ARL14EP* (Supplementary Table 7c). Nevertheless, the proximity of the fine-mapped SNPs to *FSHB* and its transcription start site, and a chromatin interaction between *FSHB* and the fine-mapped credible set (figure 5) resulted in slightly lower scaled XGBoost scores for *FSHB (0.865)* than for *ARL14EP (1.0)*. However, the significantly higher convergence score for FSHB results in a final FLAMES score of 0.075, making it the most likely effector gene of the locus. These results highlight how the FLAMES framework can correctly weigh SNP-to-gene-based and convergence-based evidence to resolve complex loci.

**Figure 5.**
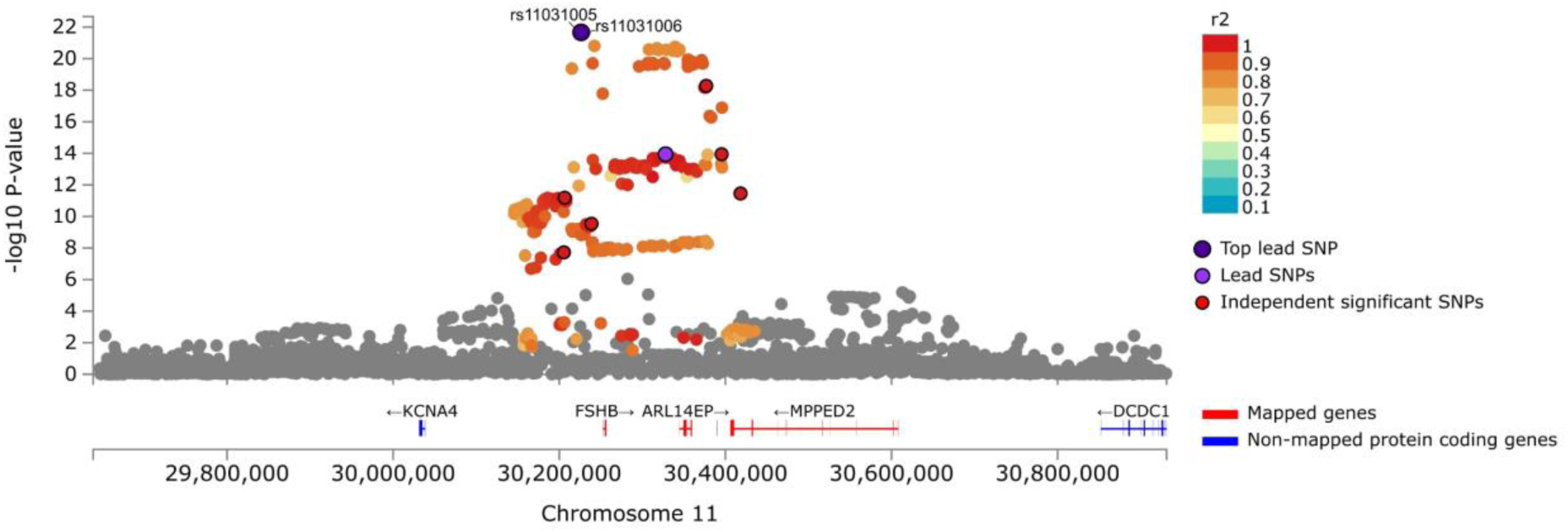
FLAMES mapped genes of the twinning FSHB locus. Locus plot showing only genes mapped by FLAMES to the credible set. The two SNPs with a PIP > 0.1 are indicated by rsID and are not clearly separated by genomic distance on this scale. Lead SNP and fine-mapping centroid are closest to FSHB although most significant variants within a gene body map to ARL14EP.

### FLAMES implicates novel temporally different gene-sets in schizophrenia and converges GWAS with ultra-rare variant analysis

Next we used FLAMES to predict the effector genes from the fine-mapping results of the most recent meta-analysis of schizophrenia^18^. We made 186 effector gene predictions, prioritizing 183 unique genes, using FLAMES after considering genes within 750kb window of the credible set centroid and using a FLAMES score cutoff threshold of 0.05 (Supplementary Table 8). Of these 183 genes, 45 were mapped to synaptic processes by SYNGO^19^ (Supplementary Table 9). The original PGC publication prioritized a list of 106 genes based on the same fine-mapping data, utilizing a custom made prioritization strategy that is tailored to prediction of risk genes for disorders affecting the central nervous system (CNS) (Supplementary Table 10). Of these 106 genes only 21 were mapped to synaptic processes by SYNGO (Supplementary Table 11). FLAMES allows for a significant increase in risk gene yield from the 255 fine-mapped schizophrenia-associated loci (p = 0.0011, one-sided Fisher exact test), whilst prioritizing a similar to larger proportion (OR = 1.32, CI95%[0.74, 2.37]) of synaptic genes with the phenotype. Given that we expect that a substantial part of disease risk and protective effects will be mediated by synaptic biology and the PGC prioritization strategy was tailored to disorders of the CNS, it appears that FLAMES predictions are accurately calibrated to take the biological context of genes into account just as well as expert-guided custom methods would. These findings suggest that FLAMES predictions recover more of the disease-relevant biology than custom prioritization methods and provide an easier and quicker path to high-confidence risk genes.

In order to perform gene ontology (GO) enrichment analysis we reran FLAMES with an adjusted version of PoPS, where the input features exclude pathway information. Running pathway analysis on the FLAMES prioritized genes using the full PoPS set might results in an increase in false positives. This pathway-naïve FLAMES analysis yields 182 predicted schizophrenia risk genes on which we performed SYNGO gene ontology enrichment analysis (Supplementary Table 12-13). Enrichment analysis was performed in a logistic regression framework, where we tested gene set enrichment conditioned on genes with high brain expression. This resulted in novel significant GO terms (Supplementary Tables 14-15). We found associations of both pre- and post-synaptic locations with the FLAMES prioritized genes (fig 6a). Notably, we found strong enrichments in modulation of post-membrane potential, postsynaptic modulation of chemical synaptic transmission and synaptic adhesion (Figure 6b). Synaptic adhesion was not observed to be associated with schizophrenia using MAGMA analysis on the SYNGO gene sets after correction for multiple testing nor was it significant when performing SYNGO analysis on the PGC defined set of risk genes (Sup. Table 14), highlighting the benefit of high quality causal gene predictions for gene set enrichment analysis.

**Figure 6.**
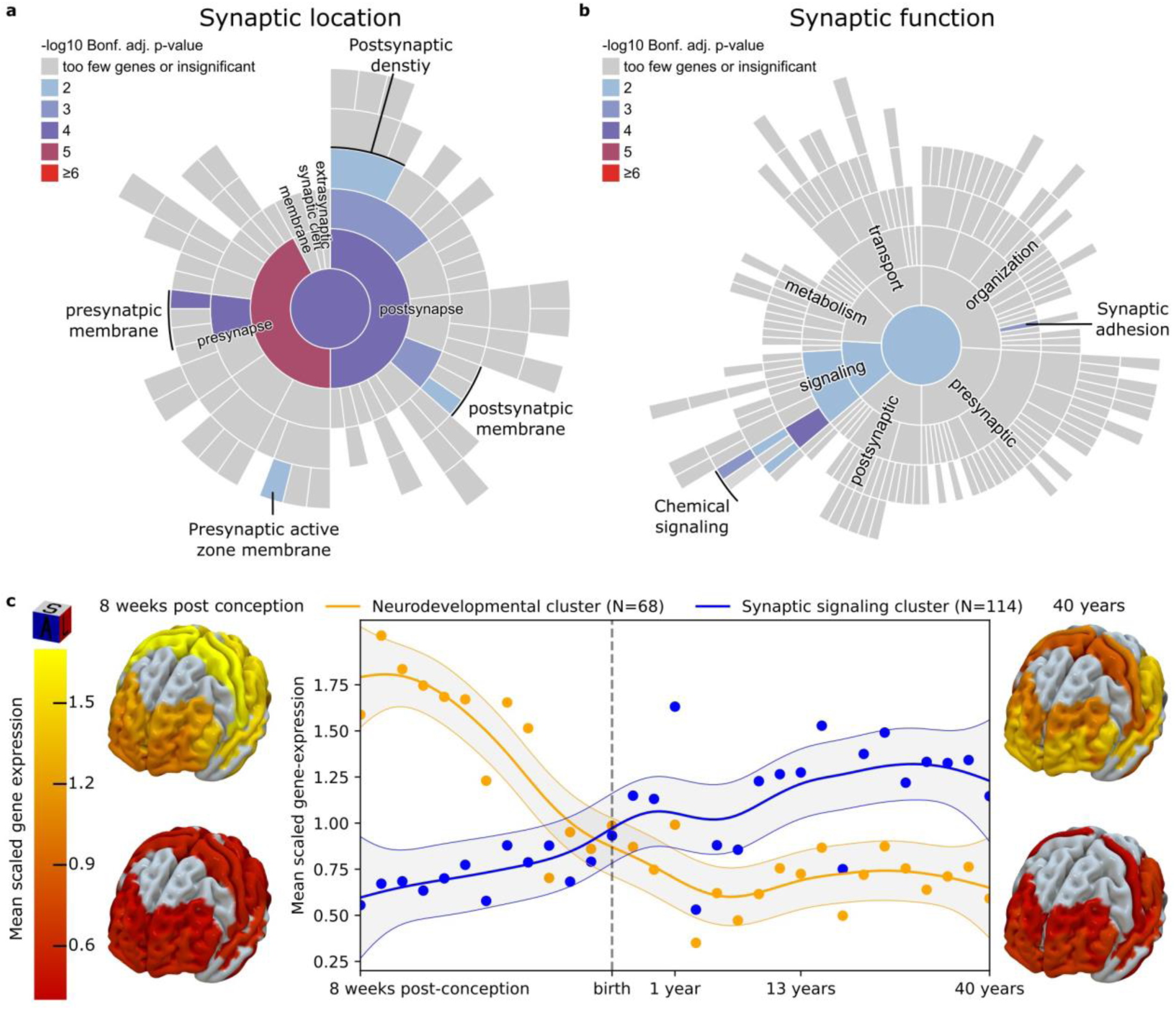
Functionality and temporal expression patterns of schizophrenia risk genes. ** a,b** SYNGO sunburst plot of synaptic cellular location and biological process GO terms respectively. Reported terms are Bonferroni-adjusted p-values of GO enrichment analysis conditioned on brain-expressed genes (methods). All significant go-terms are color coded based on their significance bin. **c**, Expression of mean-scaled schizophrenia risk gene co-expression clusters throughout the lifespan. Clusters were formed using k-means clustering on gene expression levels of schizophrenia risk genes in cortical tissue of 31 donors throughout the lifespan^20^. Gene expression per timepoint is visualized for each cluster in the brain-heatmap, grey areas represent brain regions without expression data available.

To gain further insight into the function of the FLAMES predicted schizophrenia risk genes, we extracted cortical expression of our FLAMES prioritized schizophrenia risk genes across cortical brain regions for 31 different developmental time points, similar to van der Meer *et al.* (2023). K-means clustering was performed to find gene co-expression clusters that behave similarly throughout the lifespan. Silhouette score analysis revealed that two clusters were the optimal number of clusters in the data^21,22^ (Supplementary Table 16-17). We extrapolated the expression profile across the lifespan by general additive modeling and observed that the two clusters have a distinct temporal expression profile, intersecting around birth, confirming previous findings^23^ (Fig. 6c). This suggests that these two clusters of schizophrenia risk genes play distinct roles pre- and postnatally. We found that repeated clustering of all brain expressed genes also yields two clusters as the optimal separation of expression data, with the two clusters having a similar expression pattern to the schizophrenia clusters (Supplementary Fig. 3). This indicates that these expression patterns are not unique to schizophrenia, but rather the expression patterns of schizophrenia genes can be subdivided into two major brain expression programs. To identify the biological function of each separate cluster, we performed SYNGO and MSigDB GO analysis on the genes within each cluster in a regression framework, conditioning on brain-expressed-genes (Supplementary Table 18-21). The cluster with high prenatal expression is notably enriched in synaptic adhesion and multiple neurodevelopmental gene sets, such as generation of neurons, neurogenesis, development of the striatum and development of the subpallium. The cluster with high postnatal expression is notably enriched in synaptic signaling and ion channel-related pathways. We visualized gene expression of each cluster at the earliest and latest measured time points by projecting the mean-normalized gene expression values of each cortical area onto the 3D MRI Brodmann atlas^24^. Mean cortical expression between the two clusters is significantly different (p<0.001, Mann-Whitney U test) at both 8 weeks post conception and 40 years of age. The observed increases in expression are generally cortex-wide, rather than strongly region-specific (Fig. 3c). Overall, we found that a large part of schizophrenia risk genes is active prenatally throughout the cortex, with mostly neurodevelopmental functions, and that the rest of the schizophrenia risk genes is active throughout the cortex mostly after birth and strongly enriched in functions related to synaptic signaling.

Lastly, we find that FLAMES prioritizes *GRIN2A*, *SP4*, *STAG1* and *FAM120A*, which were found to be associated with schizophrenia in rare variant analysis^25^. The PGC prioritized genes contain *GRIN2A* and *SP4*, *GRIN2A* and *SP4*, but *FAM120A* and *STAG1* did not meet the threshold for prioritization (Supplementary Table 22). This suggests that FLAMES performed better at finding convergence of rare and common signal for schizophrenia risk genes. Specifically, FLAMES implicated synaptic adhesion as a key mechanism in genetic risk burden of schizophrenia, a finding not previously reported from GWAS results. Combined, these results suggest that FLAMES can prioritize disease relevant genes, even for complex psychiatric traits.

## DISCUSSION

As recent GWAS of polygenic traits have identified hundreds of associated loci, individual investigation of each trait-associated locus to prioritize genes for follow-up study becomes infeasible. Moreover, with the increasing availability of relevant SNP-to-gene evidence, it is becoming more complex to correctly assess the relative importance of each annotation. We find that data-driven approaches for the evaluation of the relevance of SNP-to-gene annotations eliminates biases that are present in expert curated set. We show that combining SNP-to-gene-based and convergence-based methods improves prediction. We also showed that FLAMES can accurately predict the effector gene of GWAS loci.

The recent GWAS of being the mother of dizygotic twins (MoDZT) resulted in four reproducible loci which reach SNP-level genome-wide significance, with some obvious candidate genes within that locus. However, a clear link between the associated SNPs and some of these genes had so far been missing. This has been especially striking for the locus harboring the *FSHB* gene, a subunit for follicle stimulating hormone, where the most functional data prioritizes *ARL14EP*, which has no obvious biological relation to MoDZT. Here we find that FLAMES does prioritize FSHB when SNP-to-gene evidence is combined with convergence based evidence, where just SNP-to-gene evidence prioritizes ARL14EP. We speculate that the causal SNP in this credible set might affect *ARL14EP* as well, but that the regulatory effect on *ARL14EP* occurs in tissues/cell-types that are not relevant to MoDZT or that an aberrant splice isoform of *ARL14EP,* which is reverse complementary to *FSHB,* might directly impact the gene regulation of *FSHB*.

The results of FLAMES regarding schizophrenia are striking. Although FLAMES is trained to be trait agnostic, FLAMES prioritized more synaptic genes from schizophrenia GWAS results than the custom gene prioritization method used in the original schizophrenia publication which leveraged brain-specific data. This suggests that the combined SNP-to-gene and convergence approach allows for prediction of genes active in trait-relevant tissues. FLAMES predictions converged with all rare coding variants for which there were overlapping GWAS loci which indicates that FLAMES can predict known disease relevant genes even for complex traits such as schizophrenia. We find enrichment for signaling related synapse pathways that are relevant for disease risk later in life. Interestingly, we find additional evidence for the role for synaptic adhesion proteins, beyond *NRXN1* and its associated neuroligins. Whereas a role for *NRXN1* and the *CAM* pathway in schizophrenia has been previously shown^26,27^, little is known about the broader role of synaptic adhesion in schizophrenia. Our findings suggest that the genes involved in synaptic adhesion mediate schizophrenia disease risk in tandem with brain development prenatally and interfere in synaptic signaling processes postnatally.

There are some limitations to this study. First of all, there was limited weighting of SNP-to-gene interactions based on tissue type in the inputs for the XGB classifier. Broader weighting of relevant tissue might yield more accurate results. We were only able to perform tissue-specific annotation weighting in the GTEx data because it is the only dataset that has uniformly processed data across different tissue types with available expression data that was included in annotation. Secondly, it was necessary to uniformly compare tools for benchmarking but these tools that have different core assumptions so our comparison may not compare each tool in its optimal scenario. For example, FLAMES was not made to process the fine-mapping data in multiple smaller chunks, as was done for the fine-mapping for the nine-traits benchmarking set. The performance on the nine phenotype benchmarking set will most likely improve if the entire locus was fine-mapped as one. Nevertheless, we believe we selected the fairest benchmarking scenarios. Further rationale for the benchmarking scenarios can be found in the Supplementary Note. Lastly, FLAMES ultimately relies on fine-mapping the associated variants in a locus down to a credible set to reduce prediction noise. Better fine-mapping will lead to better predictions. We urge potential users to be careful when using fine-mapping output created by using out-of-sample reference panels^28^.

In conclusion, FLAMES is an integrative framework that can predict the effector gene of a GWAS locus from summary statistics and statistical fine-mapping data with high accuracy. Transitioning from associations between traits and variants to associations between traits and genes is essential for gaining a deeper understanding of the underlying biological mechanisms, aiding the translation of GWAS into meaningful functional discoveries.

## METHODS

### Conceptual framework FLAMES

The FLAMES framework is a classifier developed to predict effector genes in GWAS loci. Specifically, it combines GWAS-wide convergence of genes and locus-based biological data that links effect SNPs to genes. A prediction is made for each fine-mapped credible set in a locus. The underlying assumption is that each causal SNP will have a single gene which most strongly mediates the effect on the measured trait. Gene convergence scores are calculated using PoPS. PoPS estimates the importance of pathway, protein-protein interaction and co-expression features by regressing them on the MAGMA Z-scores of the GWAS of interest. The final PoPS score is then calculated as the sum of the beta’s of the relevant converging features. Locus-based SNP-to-gene scores are derived from an XGBoost classifier^7^ which takes fine-mapped credible set based gene annotations as an input. The inputs for the classifier are created with FLAMES Annotate, which annotates a credible-set with various SNP-to-gene annotations. The convergence based scores and the locus-based SNP-to-gene scores are scaled and combined into a final FLAMES score for each gene in the locus. There is always a single positive FLAMES score in the locus. Higher FLAMES scores denote more certainty that this gene is indeed the effector gene of the fine-mapped credible set for the studied trait. Below we will describe how credible sets are annotated, scored by XGBoost and integrated with PoPS predictions. We will then describe how the XGBoost classifier was trained and how the entire FLAMES framework was benchmarked and applied in this study.

### FLAMES annotation

FLAMES Annotate is a tool which takes as input credible sets as created by any statistical fine-mapping method that creates credible sets. Each credible set, given as a list of SNPs which contain a single causal SNP and a list with the corresponding probabilities of each of these SNPs being that causal SNP is annotated. This annotation creates annotation scores for each score in the locus based on SNP-to-gene evidence from SNPS in the credible set. There are two different types of SNP-to-gene linking annotation, either SNP-specific (e.g. fine-mapped gene expression QTLs), or regional (e.g. enhancer regions for a gene). Scores are created by multiplying the posterior inclusion probabilities (PIPs) for SNP-based annotations, or by multiplying the PIP of the fine-mapped GWAS SNP with the annotation metric of the regional annotations. The annotation metrics used can be found in Sup. Table 23. To encode locus context, relative scores are calculated by linearly scaling the maximum and minimum scores within that annotation so that the gene with the highest annotation score has a score of 1 and the gene with the lowest score has a score of 0. To generalize:

Let:

*S_g_max* be the maximum annotation score for gene *g*. *S_g_mean* be the mean annotation score for gene *g*.

*n* is still the total number of SNPs in the set.

*SNP_i_* represents the *i*-th SNP in the set.

*PIP_i_* is the posterior inclusion probability for the *i*-th SNP.

*AM_i,g_* is the annotation metric of the SNP-to-gene annotation linking *SNP_i_* to gene *g*.

Then:

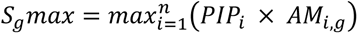

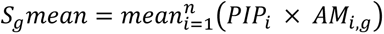

This results in an annotation matrix of genes and annotation scores. Lastly, MAGMA-Z scores, PoPS scores and locus metadata are added to the annotation matrix. The locus metadata consist of the number of genes within 50/100/250kb of the lead fine-mapped variant, the number of SNPs in the credible set, the highest PIP in the locus and the number of genes in the locus. The window of genes annotated to a locus can be specified, as well as the specific genes that are annotated to that locus. The default and the annotations on which was trained are all genes within 750kb of the fine-mapped lead SNP.

### FLAMES gene scoring

For each credible set in a locus, FLAMES outputs a predicted effector gene for that credible set. This score is calculated as follows: First XGBoost and PoPS scores are linearly scaled within the locus, so that the highest score is 1 and the lowest score is 0. Then, with the weights for XGBoost and PoPS as obtained from the calibration step, the scores are linearly combined as follows:

Let:

*C_g_* be the combined prediction score for gene *g*.

*X_g_* be the locus scaled XGBoost prediction score for gene *g* being the effector gene in the locus.

*P_g_* be the locus scaled PoPS score for gene *g*.

Then:

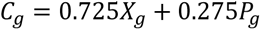

The combined scores are subsequently scaled based on the difference between the first and second highest score in the locus as given as:

Let:

*F_g_* be the FLAMES score of gene *g*.

*C_g_* be the combined PoPS and XGBoost score of gene *g*.

*C_1_* be the highest combined PoPS and XGBoost score in the locus.

*C_2_* be the second highest combined PoPS and XGBoost score in the locus. Then:

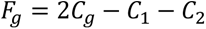

This results in a single positive score per locus, which is the FLAMES score with the highest likelihood of being the effector gene. The combination of PoPS and XGBoost scores was calibrated by a 50-fold cross validation strategy described in the section *“Training of FLAMES framework”* below.

### Creation of a data-driven training set of locus-gene pairs

To train a machine learning model we required a baseline set of GWAS loci and their true effector genes. In order to create this dataset, we linked GWAS loci of traits from the UK biobank (UKBB) with genes that have deleterious coding evidence. Genomic risk loci of these traits were created using FUMA^29^. We annotated a gene as causal for a locus if it has either significant associated missense or predicted loss of function (pLoF) variants or is implicated by gene burden tests containing at least one pLoF or missense variant in the exome wide association (ExWAS) study of that same phenotype. ExWAS results were extracted from GeneBass^11^.

Given that two nearby genes can be independently relevant for a trait and be implicated separately by GWAS and ExWAS, we aimed to reduce the amount of false positive locus-gene pairs by restricting the maximum distance between the fine-mapped GWAS signal in a locus and assigned effector genes. To do this, fine-mapping of loci was performed using FINEMAP (version 1.4.1). We created a reference panel with an LD 100,000 from unrelated UKBB individuals. FINEMAP was performed with a maximum number of causal SNPs (*k)* = 1. This threshold was chosen to increase the likelihood of predicting actual causal SNPs and minimize the number of SNP-gene pairs that can be made in a single locus for a single trait, to avoid overfitting. Each fine-mapped credible set was transformed into a 95% credible set, meaning that the credible is reduced to the smallest number of variants that together have a summed prior inclusion probability (PIP) of at least 0.95. For each fine-mapped credible set, a weighted centroid was created using the posterior inclusion probability of each SNP in that credible set. We retained all locus-gene pairs whose effector gene was no further than 750,000 base pairs of the centroid of the credible set.

### Training of FLAMES framework

We applied FLAMES annotate to 1181 loci. These 1181 GWAS loci contain a single gene which is implicated by missense/pLoF variants in an ExWAS for the same trait. For each set of maximum and summed scores for a given annotation, we retained the score with the highest odds-ratio between causal and non-causal genes. These retained scores were combined with MAGMA Z-scores and locus meta data and subsequently used in training an XGBoost classifier that aims to predict if a gene is an effector gene based on the SNP-to-gene annotations. We used a 100 iteration 5-fold cross validation randomized grid search to find the optimum parameters for our model. We subsequently used 50-fold cross validation to optimize the linear integration of PoPS and XGBoost. The data was randomly split into 50 equal parts and for each fold the model was retrained on all splits except for a single holdout split. Predictions with the retrained model were made with a different linear integration coefficient between PoPS and XGBoost ranging from 0 to 1 in 0.025 intervals on the holdout split. For each fold the integration coefficient which resulted in the highest harmonious mean of precision and recall (f1-score) was retained and the median value across all integration coefficients (0.725 FLAMES, 0.375 PoPS) was used as to integrate the XGBoost and PoPS scores.

### Benchmarking of FLAMES

We benchmarked the performance of FLAMES on the holdout validation set, high-confidence locus-gene pairs from three molecular traits, high-confidence locus-gene pairs from ExWAS data, and an expert-curated locus-gene pair dataset. A more extensive overview of the benchmarking can be found in the supplementary note.

#### Three molecular traits benchmark

In the three molecular traits datasets we benchmark the prediction of FLAMES on loci from traits with easily interpretable loci given extensive domain knowledge of these traits. We created loci using FUMA from the summary statistics and fine-mapped the loci using FINEMAP and a reference panel of 100,000 unrelated individuals from the UKBB. Fine-mapping was performed with a maximum number of causal SNPs (*k)* = 10. Each individual credible set was mapped using FLAMES on the default settings. In this specific benchmark, we were interested if the actual causal gene would be recovered at least once given that all credible sets in the locus could be used for prediction. We therefore define recall in this dataset as the number of recovered unique genes divided by the number of total unique genes. We extracted L2G results from the L2G database if they make a prediction for the same locus. To avoid annotation biases we only compare genes that are scored by both methods, and remove genes that are only scored by a single method. We calculated cS2G scores by multiplying the cS2G scores for a gene with the PIPs of each SNP and sum the resulting scores per gene. Genes not mapped by L2G or FLAMES were dropped, and genes not mapped by cS2G but mapped by the other methods received a score of 0.

#### Expert curated locus-gene-pairs benchmark

We extracted the L2G scores and fine-mapping results for all loci for which full summary statistics were available from the Open Targets platform. We annotated these credible sets with FLAMES annotate to ensure that the benchmarking is done on the exact same credible sets. We extracted all credible sets from the original L2G training set for which full summary statistics were available from the L2G platform, along with the corresponding L2G predictions. FLAMES was run with default settings. We calculated performance metrics on genes annotated by both methods to eliminate any biases in annotation. An example of this: these loci were selected to have their effector gene within 500kb, which is the maximum range in which L2G maps genes, whereas FLAMES maps up to 750kb by default.

#### Nine phenotypes with ExWAS-implicated genes benchmark

When benchmarking the expert curated dataset, we compare the predictions of single credible sets to each other, given that Ei bins all the credible sets in a locus together and only has a single locus with multiple true-positive genes. Comparisons between L2G, Ei and FLAMES are always performed on the genes which were scored by all three methods. FLAMES was performed on the fine-mapping results as performed by Forgetta et al in the original Ei paper^2^. This fine-mapping was performed by splitting loci into 250kb chunks and fine-mapping with multiple causal variants. We therefore transformed each credible set so that the PIP of each SNP is multiplied by its Bayes factor and divided by the sum of Bayes factors of the entire credible set. This results in a credible set that is akin to the results when fine-mapping for a single causal variant in the locus. Ei scores were acquired from the original publication ^5^, L2G scores were acquired from the L2G database, extracting each locus that maps the true-positive gene for the corresponding GWAS. L2G and FLAMES predictions for each credible sets were averaged within each locus. We calculated cS2G scores by multiplying the cS2G scores for a gene with the PIPs of each SNP and sum the resulting scores per gene. Genes not mapped by L2G or FLAMES were dropped, and genes not mapped by cS2G but mapped by the other methods received a score of 0.

### Calibration of FLAMES

To calibrate FLAMES scores we aimed to create a benchmarking set that contained the least amount of biases. We therefore merged the bechmarking set with ExWAS implicated genes of nine phenotypes and the set with genes from domain knowledge of three molecular traits, as no prior SNP to gene knowledge was used to curate these datasets. We incrementally lowered the FLAMES score threshold from 1 to 0 in increments of 0.05 and retained the score with at least 75% precision.

#### Application of FLAMES to Mother of Dizygotic Twins GWAS

Given that the a large portion of the MoDZT controls come from the UKBB, and that the sample population is European, we created an LD reference panel based on 100,000 unrelated individuals from the UKBB for fine-mapping. Given that we have to use a reference panel that is not a precise subset of all the data across cohorts we restrict maximum number of causal SNPs per locus (*k)* = 1. FLAMES was ran with default settings (max distance to gene = 750kb, only protein coding genes annotated).

#### Application of FLAMES to Schizophrenia GWAS

We ran FLAMES with default settings (max distance to gene = 750kb, only protein coding genes annotated) on the fine-mapping output as reported in in the PGC-3 schizophrenia fine-mapping study^18^. We additionally ran FLAMES with a pathway naïve version of PoPS with which we prioritized genes that were eligible for pathway analysis. We subsequently extracted the FPKM normalized gene expression values across the lifespan of the pathway naïve FLAMES prioritized genes from all cortical tissues in the Brainspan data. Expression per gene was normalized across timepoints, by dividing the expression levels of each by the mean expression of that gene across all timepoints similar to van der Meer *et al.* (2023). We subsequently clustered the data by k-means clustering, with the optimum number of clusters being determined by selecting the number of clusters with the highest average silhouette score. Gene ontology (GO) enrichment analysis of the biological processes and cellular component ontologies was performed on the SYNGO (release 1.2, 2023-12-01) and MSigDB (v2023.1) GO datasets. Enrichment analysis was performed using a logistic regression framework, in which the genes in the gene set are tested competitively against the genes outside of the gene set by regressing them onto the FLAMES predictions. An additional conditional analysis was performed to verify if the enrichment was independent of the general brain-related signal, by repeating the regression analysis including a binary vector indicating if genes are expressed in the brain as a covariate. This list of brain-expressed genes was previously defined by the SYNGO consortium and can be downloaded from the SYNGO platform^19^. In order to create the cortical maps, we have manually matched the ontologies between the modified Brodmann atlas of the Brainspan project in two stages of development - 8 weeks post conception and adult brain (40 years old) - and the 3D MRI Brodmann atlas by Pijnenburg et al (2021)^24^. The matching can be found in Supplementary Table 24. Only cortical areas were included in the plot for consistency of atlas usage. Transient brain structures were not included in the plot.

## DATA AND CODE AVAILABILITY

The code for installing and running FLAMES can be accessed via https://github.com/Marijn-Schipper/FLAMES.

The reference data needed for running FLAMES and/or running (pathway-naïve) PoPS can be found here: https://zenodo.org/uploads/10409723

The data and code used for the analyses in this paper and creation of the figures can be accessed via https://github.com/Marijn-Schipper/FLAMES_paper_analyses

## ACKNOWLEDGEMENTS

We thank M. de Hemptinne, K. Heilbron and the members of the Complex Trait Genetics lab for their input and discussions. This project was funded by NWO Gravitation: BRAINSCAPES: A Roadmap from Neurogenetics to Neurobiology (Grant No. 024.004.012). This research has been conducted using the UK Biobank Resource under Application Number 16406.

