## Supplementary notes for "Gene prioritization in GWAS loci using multimodal evidence"

Benchmarking of gene prioritization methods

Comparing the different methods is difficult since many choices affect comparative performance metrics, often without affecting the predictive power of the method. Moreover, some methods have different assumptions, inputs or goals. E.g. Ei tries predict for all genes mapped to locus whether they are causal, giving the possibility to rank them, whereas L2G and FLAMES make a prediction per credible set. L2G does allow multiple predictions but rarely so, and it is suggested to take the highest-scoring gene, whereas FLAMES inherently makes a single prediction per credible set. Here we outline the choices made in the different benchmarks as displayed in Figure X.

*Benchmarking three molecular traits*

We wanted to benchmark all available methods on high-confidence locus-gene pairs. The aim of this benchmark is to see whether the benchmarked methods can map credible sets within each locus to one of the locus gene pairs. Given that multiple genes can be linked to a locus, we also see how many genes are recovered by each method. It is possible that the GWAS used contains to actual biologically relevant credible sets for a given gene, meaning that a non-100 % recall might be a result of underlying biology rather than failing of a method. To compare the performance of cS2G, FLAMES, L2G and PoPS we assume the following:

cS2G, FLAMES and L2G require fine-mapping results to make a single prediction per credible set. The PoPS is recommendation is to take the highest scoring gene in the locus. To capture the performance over the same genomic region we take the results for the different methods as follows:

cS2G: We annotate all the fine-mapped credible sets within the relevant FUMA loci with PIP weighted L2G scores.

FLAMES: We run FLAMES on all the fine-mapped credible sets within the relevant FUMA loci.

L2G: We extract the L2G results belonging to credible sets for which the GWAS lead SNPs fall within the FUMA defined loci.

PoPS: We take the gene with the highest PoPS score within each GWAS locus.

Closest gene: For each credible set, we take the closest gene by PIP weighted centroid to the gene body.

*Benchmarking 9-phenotype benchmark*

Ei is different to L2G, FLAMES and cS2G as the entire unit of analysis shifts from single SNP/credible set to the entire locus. This means that we need to convert L2G and FLAMES scores to scores for all the credible sets within each locus. The difficulty here is that L2G and FLAMES scores do not incorporate the fine-mapping relevance of the credible sets, only the confidence of a gene being the effector gene of a credible set. With the unit of analysis shifting to larger loci, it matters which genes are annotated to the locus to compare performance in an Area Under the Precision-Recall curve (auprc), as this can be strongly skewed by class imbalance. We solve this by taking an inner merge of the predictions by the methods that annotate multiple genes in a locus based on location. We extract the results as follows:

cS2G: We annotate all the fine-mapped on the credible set created by re-scaling the merged fine-mapping results from Forgetta et al.

Ei: The Ei predictions from Forgetta et al.

FLAMES: We run FLAMES on all the fine-mapped on the credible set created by re-scaling the merged fine-mapping results from Forgetta et al.

L2G: We extract the L2G scores for each locus that has a score for any of the true positive genes from the 9-phenotype benchmark set.

PoPS: We take the gene with the highest score from each locus.

We take the inner join of gene scores from Ei, FLAMES and L2G, effectively eliminating different gene-locus annotations. We note that L2G and Ei are inherently incompatible here, but FLAMES can take a custom mapping of candidate genes for a given locus. In the benchmark from Forgetta et al, the highest scores are retained for each gene. We think this is relatively unfair, as only the Ei actually takes into account the betas and bayes factor from the GWAS and fine-mapping. The scores of the other methods only denote the certainty that a gene in the effector gene of a SNP/credible set, not how relevant the GWAS signal in this gene is. These methods assume that the finding of independent loci and fine-mapping of them has already ensured that you are only looking at relevant signals. We provide two metrics for the non-Ei methods. The results from the credible set closest to the true-positive gene, and the sum of all gene scores across credible sets.

Benchmarking of L2G set:

We extracted the results from the original L2G training loci as described in mountjoy et al. for loci that have available fine-mapping results from the Open Targets platform. To eliminate differences in fine-mapping in this benchmark, we ran cS2G and FLAMES on the fine-mapping results from Open Targets.
