## Supplementary figures for "Gene prioritization in GWAS loci using multimodal evidence"

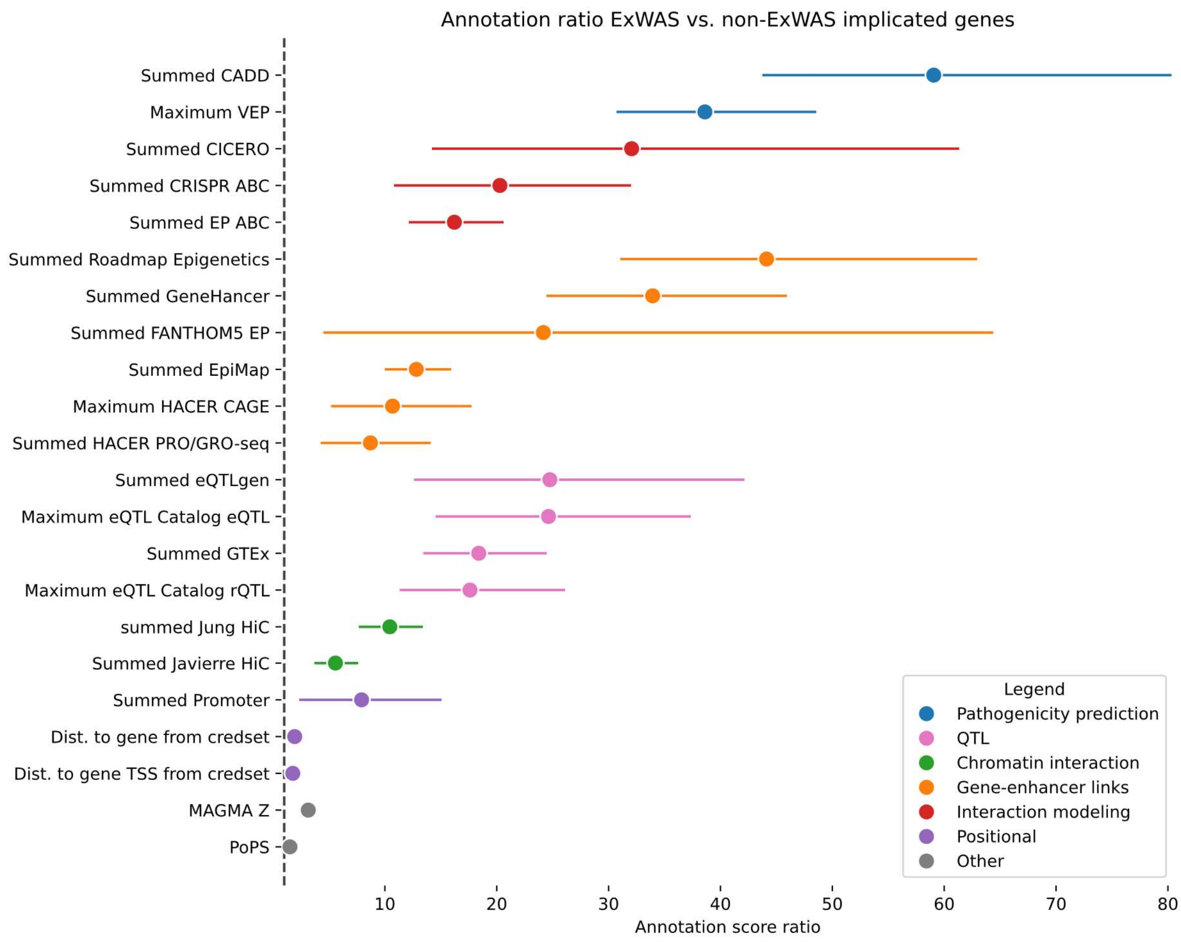


**Supp. Figure 1 | Ratio of annotation scores in L2G expert-curated causal genes.** Ratios of average annotation score per annotation in ExWAS implicated gene in GWAS locus vs. the rest of the genes in the locus. Confidence intervals were calculated by 1000 times bootstrapping.

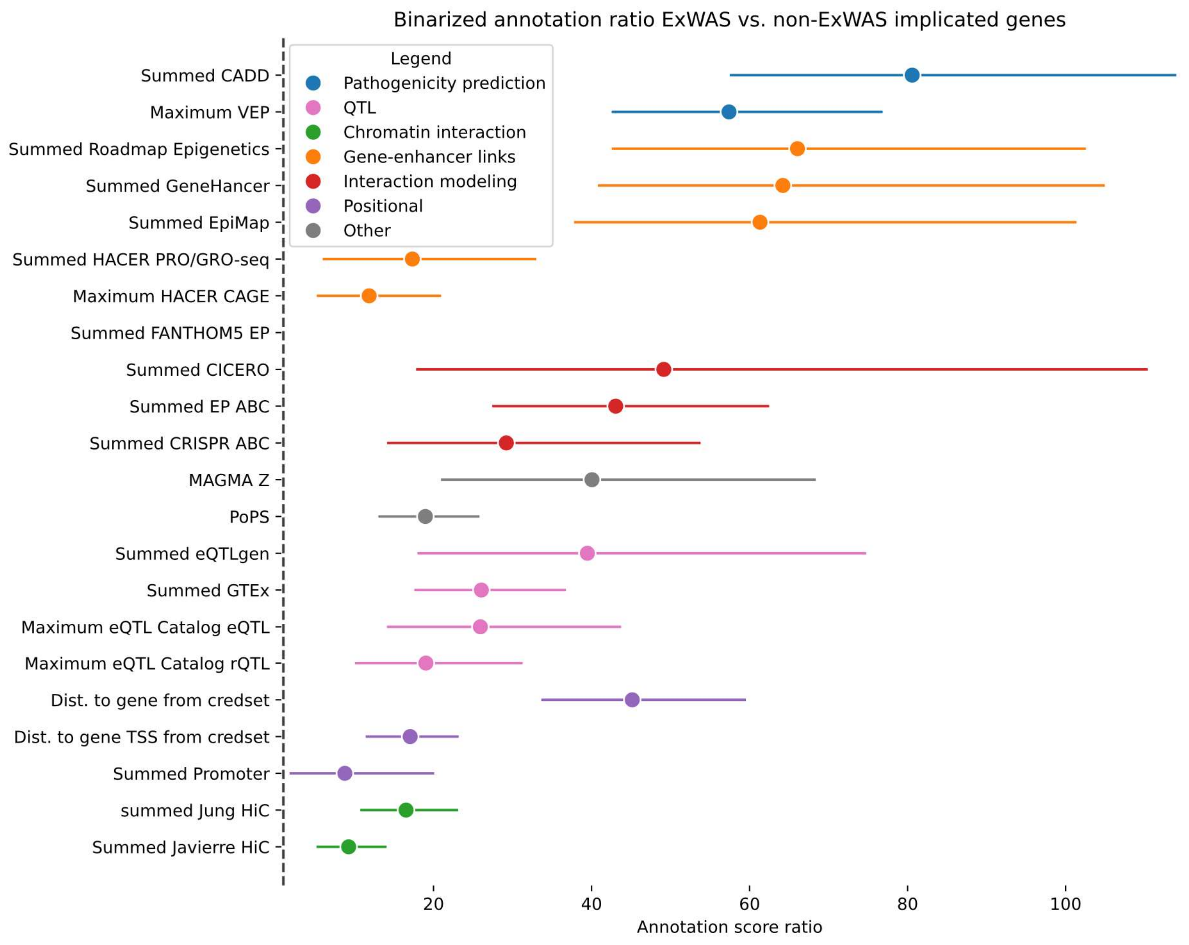


**Supp. Figure 2 | Odds ratio of expert-curated causal gene having the highest annotation scores in the locus.** Odds ratio of highest annotation score in the GWAS locus belonging to the ExWAS implicated gene in the locus. Confidence intervals were calculated using 1000 times bootstrapping.

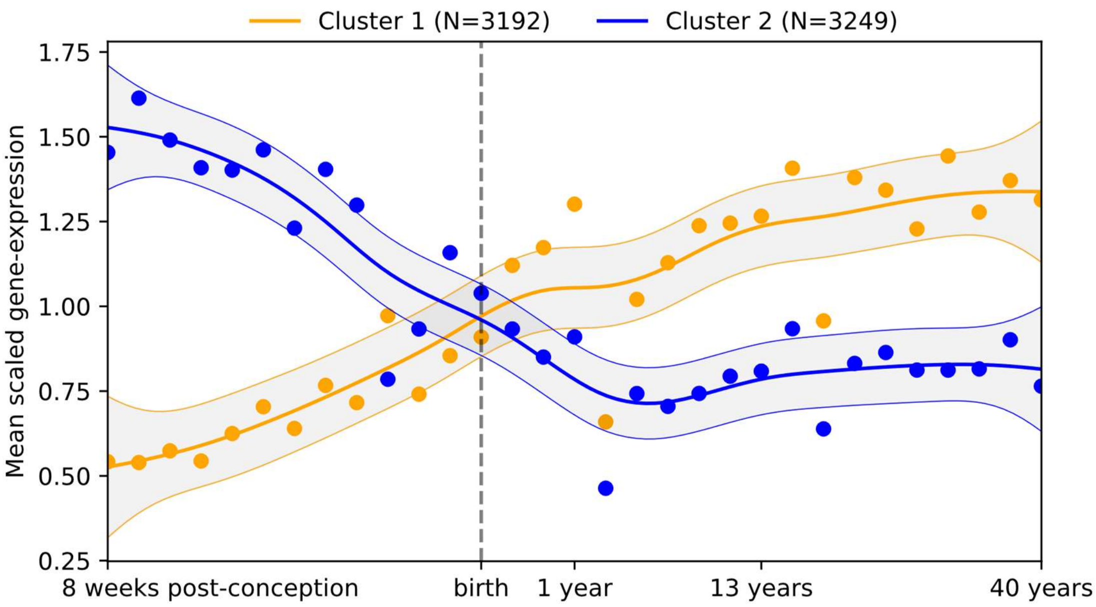


**Supp. Figure 3. Brain expressed genes expression profile in BRAINSPAN. |** Expression profile of BRAINSPAN brain expressed genes after k-means clustering. Gene expression is mean scaled per gene, and averaged across all genes in the cluster per timepoint.
